# Detection of SARS-CoV-2 in the community by nucleic acid amplification testing of saliva

**DOI:** 10.1101/2021.09.15.21263644

**Authors:** Filippo Fronza, Nelli Groff, Angela Martinelli, Beatrice Zita Passerini, Nicolò Rensi, Irene Cortelletti, Nicolò Vivori, Valentina Adami, Anna Helander, Simone Bridi, Michael Pancher, Valentina Greco, Sonia Iolanda Garritano, Elena Piffer, Lara Stefani, Veronica De Sanctis, Roberto Bertorelli, Serena Pancheri, Lucia Collini, Alessandro Quattrone, Maria Rosaria Capobianchi, Giancarlo Icardi, Guido Poli, Patrizio Caciagli, Antonio Ferro, Massimo Pizzato

**Author notes:** These authors contributed equally to the work (Filippo Fronza, Nelli Groff, Angela Martinelli, Beatrice Zita Passerini, Nicolò Rensi, Irene Cortelletti, Nicolò Vivori). Corresponding author Department of Cellular, Computational and Integrative Biology, University of Trento, Via Sommarive 9, 38123, Italy.

## Abstract

Efficient wide-scale testing for SARS-CoV-2 is crucial for monitoring the incidence of the infection in the community. The gold standard for COVID-19 diagnosis is the molecular analysis of epithelial secretions from the upper respiratory system captured by nasopharyngeal (NP) swabs, which requires the intervention of trained personnel. Given the ease of collection, saliva has been proposed as a possible substitute to support testing at the population level. Here we describe the set-up of a laboratory, in an academic context, for the high-throughput screening of SARS-CoV-2 in the saliva from the community. A novel saliva collection device was designed to favour the safe and correct acquisition of the sample as well as the processivity of the downstream molecular analysis. To test the performance of the system,1025 paired saliva and nasopharyngeal samples were collected from individuals recruited at a public drive through testing facility and analysed in parallel. An overall moderate concordance (68%) between the two tests was found, with evidence that neither test can diagnose the infection in 100% of the cases. While the two tests performed equally well in symptomatic individuals, their discordance was mainly restricted to samples from convalescent individuals. The saliva test was at least as effective as NP swabs in asymptomatic individuals recruited for contact tracing. Our study, therefore, indicates that saliva testing can be a reliable tool for wide-scale COVID-19 screening in the community.

## Introduction

SARS-CoV-2, the coronavirus causing COVID-19, has spread in all continents since the end of 2019, quickly becoming pandemic. The infection causes a respiratory syndrome which can be lethal especially in individuals affected by other pathologies and in the elderly population. The widespread diffusion of the virus has exposed the healthcare systems worldwide to an unprecedented pressure, quickly overpowering hospitals and healthcare professionals. In order to control the spread of the virus, to protect the individuals most at risk and to preserve the functionality of the healthcare system, the capability to promptly track the presence of SARS-CoV-2 in the community remains critical (Mercer & Salit, 2021).

For long-time the gold standard to reliably diagnose respiratory infections has been the analysis of secretions from the mucosa of the upper respiratory epithelium collected by nasopharyngeal swabs. For the diagnosis of SARS-CoV-2, maximum sensitivity is ensured by detecting the viral RNA in the swab using RT-PCR (Vandenberg et al., 2020). However, the success of the diagnosis relies on the correct procedure for collecting the specimen (Marty et al., 2020), with the swab that should penetrate the nasal septum and reach the epithelium of the nasopharynx to absorb respiratory secretions. This is an invasive procedure performed only by skilled healthcare workers which causes discomfort to the individuals being tested and is poorly suitable for massive scale testing in the community given the requirement of thousands of trained professionals. In addition, swab collection poses a major biohazard risk for the personnel collecting the specimens who is directly exposed to the aerosol generated by the individual being tested. For these reasons, the possibility of using saliva for COVID-19 testing has been explored soon after the virus was declared pandemic, as an alternative to nasopharyngeal swabs (To et al., 2020)

Several studies have investigated the presence of SARS-CoV-2 in oral fluids at different stages of the infection by RT-PCR, comparing the sensitivity of virus detection in saliva and in nasopharyngeal swabs. The current literature provides a consensus suggesting that viral load found in saliva is adequate for a reliable diagnosis, with most of the investigations reporting more efficient detection of SARS-CoV-2 in saliva than nasopharyngeal swabs (Kapoor et al., 2021; Tan et al., 2021; Warsi et al., 2021). Despite the encouraging evidence, adoption of widescale saliva testing remains hampered by the lack of standardization of the method of collection and the analytical procedure (Atieh et al., 2021). Available devices for collecting saliva can be very different, potentially impacting the sensitivity of detection. Popular methods include the absorption of saliva on a pad from which the fluid has to be mechanically eluted, or drooling into a test tube (Allicock et al., n.d.). In either case, the degree of viscosity, which varies among subjects, can hinder downstream processing, interfering with pipetting, causing contamination, and impacting on the efficiency of the RNA extraction.

Here we describe a diagnostic initiative implemented within an academic setting (the University of Trento) to assist the local health service with testing and screening activities for SARS-CoV-2 infection in the general population. To this end, a workflow for mass scale analysis of saliva samples was established, starting with designing a system that facilitates the autonomous and safe collection of saliva and the semi-automation of the downstream processing of samples. This strategy was tested in the context of the general population, including symptomatic and asymptomatic individuals which were subjected to both saliva and nasopharyngeal swab analysis allowing for a direct comparison.

## Results

### Design of an analytical workflow allowing large-scale screening of SARS-CoV-2 in saliva

With the scope of assisting the safe self-collection of saliva by the users and at the same time facilitating high processivity of the downstream molecular analysis, a novel saliva collection device was first designed. Standard 13 mm x 80 mm tubes already widely used for collecting nasopharyngeal swabs were equipped with an insert functioning as a straw for the deposition of saliva (Figure 1A). To prevent contamination of the exterior of the tube with oral fluid and avoid the potential dispersion of contaminated material in the environment, the insert was made to fully enter and remain locked inside the tube after saliva collection. The straw was designed to be compatible with the direct introduction of 1250 µl pipette tips for withdrawing the oral fluid, avoiding the need of further processing steps, such as centrifugation, before proceeding with the analytical protocol.

**Figure 1.**
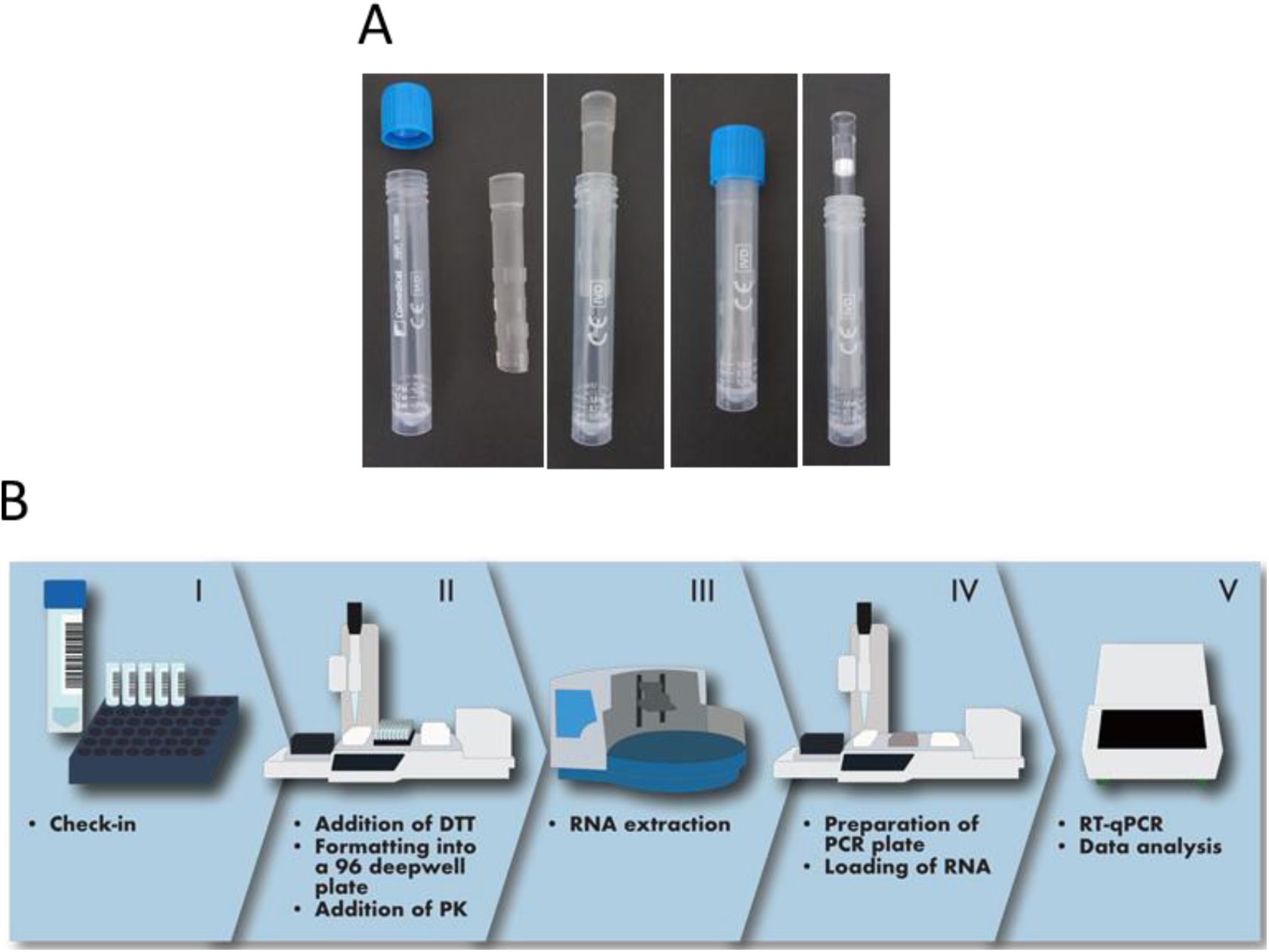
Description of the organisation of the analytical process. (A) The saliva collection device used in this study consists of a standard 13 mm tube provided with an insert to aid the deposition of saliva, which remains inside the tube without interfering with the introduction of pipette tips. (B) The analytical process begins with the tubes being prepared on racks (I) accepted by a robotised multichannel pipettor with adjustable tip spacing (II) which produces 96-well formatted plates for batched RNA extraction (III). PCR plates are then assembled with a dedicated robotised pipettor (IV) before amplification in a thermocycler (V). Transition between steps is performed by an operator.

Subjects enrolled in the study were instructed to collect between 0.5 and 1 ml of saliva into the tube, as indicated by labels on the tube itself (Figure 1A). The tubes with saliva were mounted on a rack and processed using a robot-operated multichannel pipette with adjustable tip spacing, allowing the deposition of the oral fluid into a standard 96-well formatted plate (Figure 1B). To address the potentially high viscosity of saliva, 1 ml of a 6. 5 mM DTT solution was first added to the samples using the same robotized pipette. This addition proved very effective at increasing fluidity, facilitating automatic pipetting, and preventing formation of threads of saliva on pipette tips, possibly causing cross-contamination. Based on the efficiency of extraction of an internal control added to the samples being processed, DTT-treatment was found compatible with the effective recovery of nucleic acids in every sample analysed. Saliva samples were automatically formatted into multi-well plates for downstream RNA extraction and RT-PCR, operated in batches of 96 (Figure 1B). Sample check-in required to enter the analytical line, the tracking of the samples through the different steps of the process, the evaluation of results and the final reporting to the health service repository was managed by a custom-made middleware which was instrumental for coordinating and instructing the operators through the different analytical phases. The analytical cycle, from addition of DTT to the samples to the completion of RT-PCR, lasts 3 hours with a processivity of 384 saliva samples per hour per production line.

### Global comparison of SARS-CoV-2 detection in saliva and NP swabs

Nasopharyngeal swabs and salivary samples were collected from 1025 individuals coming to a public drive-through testing facility in Trento, Italy. Nasopharyngeal swabs were administered by healthcare workers while saliva was collected autonomously by each person enrolled in the study. Among the people investigated were symptomatic and asymptomatic subjects from the general population, as well as healthcare professionals undergoing routine periodic screening (Table 1). Saliva samples and NP swabs were analysed using the same RNA extraction method and RT-PCR kit, and compared based on the detection of the RdRP gene over 40 PCR cycles. The amount of saliva collected by each subject was the only criterion adopted for excluding samples from analyses. Tubes containing visibly less than 0.5 ml of oral fluid were removed from further processing. Out of 1025 samples collected, only 22 (2.1%) were discarded based on this criterion.

**Table 1.** Characteristics of the subjects recruited in the studies.

|  | n. of individuals | Positive NP swabs |  | Positive saliva samples |  |
| --- | --- | --- | --- | --- | --- |
|  |  | n | % (95% CI) | n | % (95% CI) |
| **Total** | 1003 | 344 | 34.3 (31.4-37.3) | 312 | 31.1 (28.3-34.0) |
| **Males** | 434 | 175 | 40 (35.8-45) | 158 | 36.4 (32.0-41.0) |
| **Females** | 569 | 169 | 29.7 (26.1-33.6) | 154 | 27.1 (23.6-30.9) |
| **Contact tracing** | 193 | 56 | 29.0 (22.7-36.0) | 65 | 33.7 (27.4-40.6) |
| **Convalescents** | 442 | 333 | 75.3 (71.1-79.1) | 235 | 53.2% (48.5-57.8) |
| **Screening** | 351 | 4 | 1.1 (0.3-3.0) | 4 | 1.1 (0.3-3.0) |
| **Suspects** | 17 | 8 | 47.1 (26.2-69.0) | 8 | 47.1 (26.2-69.0) |

Age (years) mean +/- SD: 41.7 (+/-15.2)

Out of 1003 individuals tested, the number of SARS-CoV-2 positive saliva samples was 312 (31%) while SARS-CoV-2 was detected in 344 (34.3%) NP swabs (figure 2A). However, the total positive subjects detected by either the saliva or the nasopharyngeal test methods were 419 (41.8%), with 237 samples found positive and 584 samples negative with both tests, giving a total concordance of 81.8% with a kappa index of 0.588 (Figure 2A).

**Figure 2.**
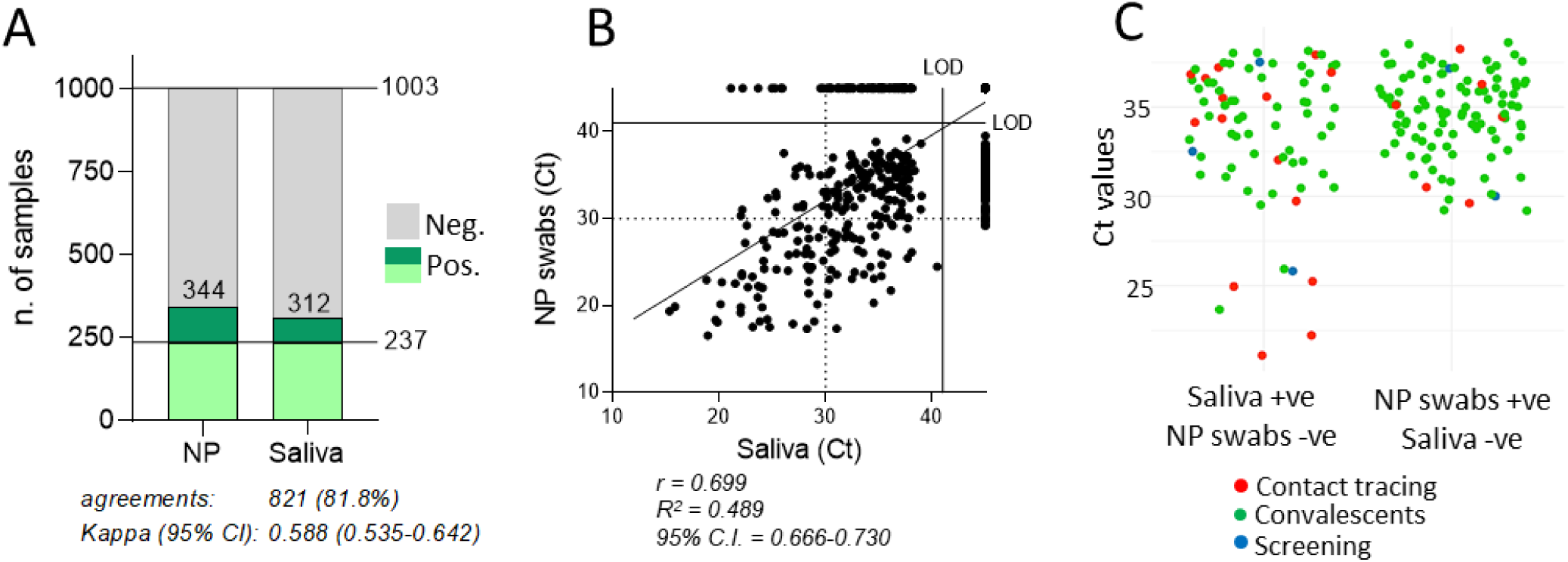
Global concordance between SARS-CoV-2 detection in saliva and NP swabs. (A) Number of individuals that tested positive (green) or negative (grey) for SARS-CoV-2 in the saliva or NP swabs. Light green: individuals that tested positive in both types of samples. The percentage of agreements between results from saliva samples and NP swabs and the kappa index with 95% confidence interval (CI) is indicated. (B) Correlation between RT-PCR cycle threshold (Ct) values of paired nasopharyngeal swabs and self-administered saliva samples (n=1003). Shown is the fitted curve of the linear regression, the Pearson correlation coefficient (r), the goodness of fit (R^2^) and the 95% confidence interval (CI). LOD: limit of detection. (C) Ct values of saliva samples and NP swabs from individuals testing positive with only one of the two methods, categorized by group of subjects as indicated by the colours specified in the legend.

Considering the NP swabs as reference, the globally sensitivity of the saliva test was 68.9% (237/344) with a specificity of 88.6% (584/659). By assessing all subjects in which SARS-CoV-2 was detected, either in the saliva or in the NP swabs, the sensitivity of detection was 74.5% (312/419) with the saliva and 82.1% (344/419) with the NP swabs, therefore indicating moderately higher sensitivity with the latter.

When Ct values obtained with the two methods were compared (Figure 2B), a good and statistically significant correlation was found (r=0.6994 with p value <0.0001), indicating a general trend with individuals with high viral loads in the NP swabs presenting also with high viral load in saliva. However, in 182 subjects, SARS-CoV-2 was discordantly detected in the two types of samples, since 75 individuals tested positive in the saliva and negative in the NP swabs, while 107 subjects tested positive in the NP swab and negative in the saliva (Figure 2).

Interestingly, while all the 107 positive NP swabs produced Ct values higher than 29, a cluster of 7 samples among the 75 positive saliva samples generated Ct values ranging between 21 and 26 (Figure 2B and 2C), irrespectively of the group of individuals being tested. Therefore, some individuals with exceptionally high viral load in the saliva failed to test positive in the NP swab.

### Saliva and NP swabs give concordant results in symptomatic subjects and in individuals recruited from contact tracing

The subjects enrolled in the study included individuals undergoing testing for having been recently in contact with infected persons (contact tracing), convalescent individuals tested to confirm their recovery, healthcare professionals for routine screening or symptomatic individuals suspected of infection (Table 1). Positive individuals recruited for contact tracing are likely to represent initial stages of the infection, in contrast to convalescent subjects which are tested after clinical resolution of the infection.

We compared the performance of saliva and NP swabs across all groups. Our study included a total of 32 individuals that presented with respiratory symptoms. Remarkably, though the sample is numerically small, both assays detected the same 15 positive individuals (making 46.9% of the total) with a concordance of 100% (Figure 3). In contrast, among 971 asymptomatic subjects, a higher number of infections were detected in NP swabs (327) than in saliva (295), with 81.26% concordance between the two methods (Figure 3). Given that the two tests performed equally among symptomatic cases, the two tests were further compared in different groups of asymptomatic subjects. Among 442 convalescent subjects, NP testing revealed 276 positive cases, 69 more than those detected in saliva (207), resulting in 64.93% concordance with a weak kappa index (0.287). Among 193 subjects recruited from contact tracing, slightly more positive cases were revealed in saliva than NP swabs (65 vs 56), with a good concordance of 89.12% and a kappa index of 0.748. This result suggests that while the oral fluid is less sensitive than NP swabs for detecting the virus during convalescence, saliva is at least as good as NP swabs during the early stages of the infection.

**Figure 3.**
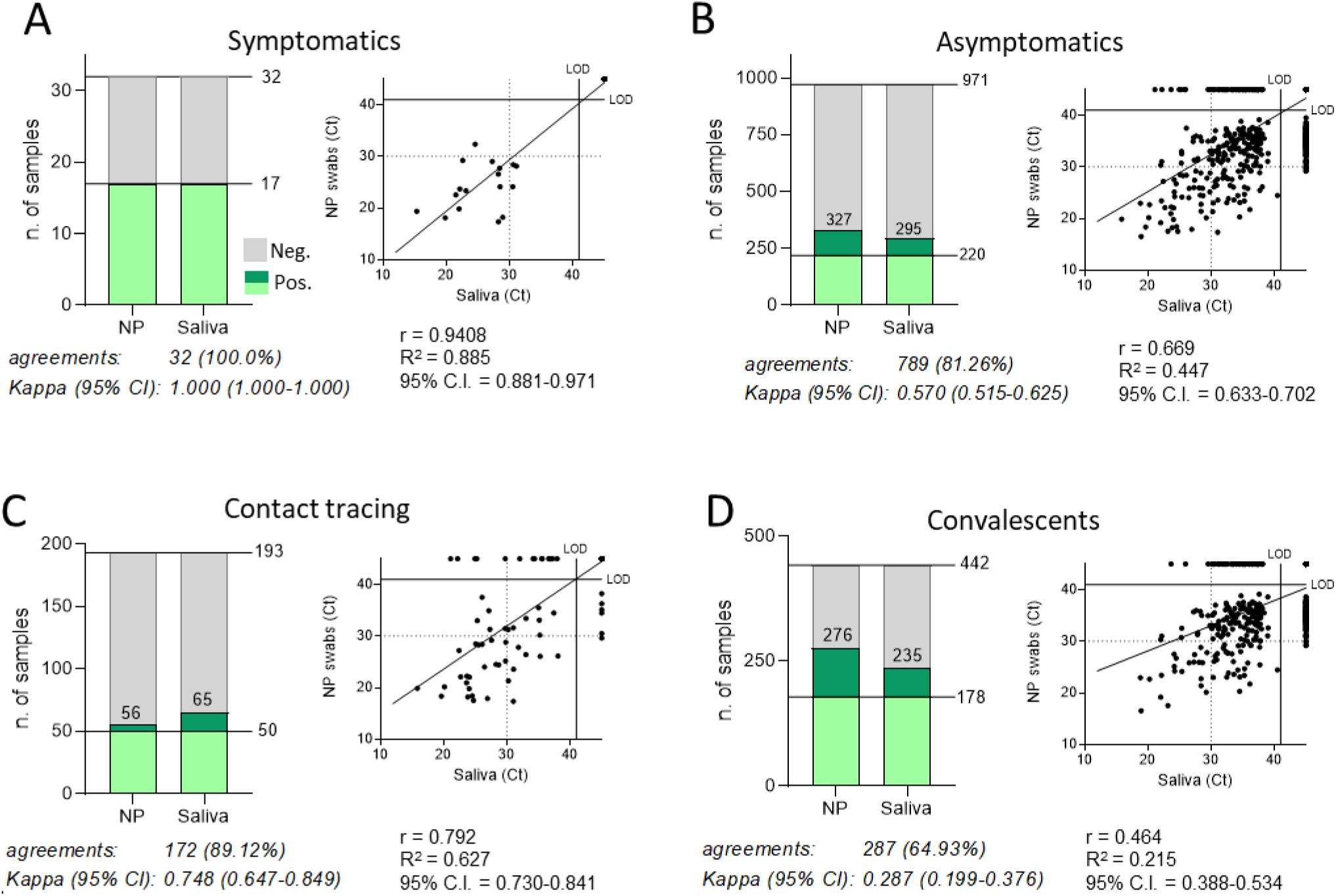
Concordance between SARS-CoV-2 detection in saliva samples and NP swabs in different groups of individuals tested. In each panel, the histograms represents the number of individuals that tested positive and negative, and dot plots the RT-PCR cycle threshold (Ct) values of the paired nasopharyngeal swabs and self-administered saliva samples. Light green: individuals that tested positive in both types of samples. The percentage of agreements between results from saliva samples and NP swabs and the kappa index with 95% confidence interval (CI) is indicated below the histograms, as well as the fitted curve of the linear regression in the dot plots, with the Pearson correlation coefficient (r), the goodness of fit (R^2^) and the 95% confidence interval (CI). LOD: limit of detection.

## Discussion

To control the spread of SARS-CoV-2, the implementation of an efficient strategy to track the virus in the population is paramount (Mercer & Salit, 2021). To this end, saliva represents an ideal tool, given that it is easily accessible and can be self-collected with non-invasive procedures without involving specialized personnel. Accordingly, our study has tested a semi-automatic workflow for analysing saliva samples from the general population. Samples were collected with a novel device designed to increase biosafety and favour highly processive molecular analysis. Semi-automation was implemented to address the need of high-throughput diagnostics, allowing reduced human intervention while keeping the analytical line flexible and compatible with reagents from different providers. At the same time the instrumentation adopted here represents a moderate financial investment compared with fully automated systems currently available on the market and can be easily converted to different applications when SARS-CoV-2 large-scale testing will no longer be required.

Our experience with this analytical process confirmed its suitability for large-scale analysis. Of note, 97.9% of samples were correctly self-collected by the subjects enrolled in the study. The parallel examination of saliva and nasopharyngeal specimens from 1003 individuals revealed discordant detection of SARS-CoV-2 in 182 subjects (Table 2), a result consistent with most data in literature, as reported by a recent review that analysed 58 studies published since the start of the pandemic (Tan et al., 2021). Such level of discordance between the two tests indicates that no approach can diagnose the infection in 100% of cases. However, stratification with different groups of individuals recruited in the study revealed interesting differences in efficiency of viral detection by the two tests. SARS-CoV-2 was identified with 100% concordance in saliva and NP swabs among symptomatic patients. A good concordance was also observed among asymptomatic individuals recruited for contact tracing, with a slightly higher number of positive samples detected in saliva than NP swabs. In contrast, poor concordance was observed in convalescent subjects with a higher number of SARS-CoV-2 positive samples detected in NP swabs than in saliva. This result suggests an inherent property of the biological samples that makes saliva an efficient matrix for the detection of the virus during the early stages of the infection and in the presence of symptoms, rather than late after infection, during convalescence.

Results obtained with this study agree with other studies that have ascertained that soon after infection SARS-CoV-2 is abundantly detected in saliva (Pan et al., 2020; To et al., 2020; Zhu et al., 2020), in line with evidence that indicates that the epithelium associated with the salivary glands is an early site of virus replication (Huang et al., 2021; Matuck et al., 2021; Pascolo et al., 2020). These results also agree with another recent investigation which confirms that SARS-CoV-2 is more readily detected in saliva rather than in NP swabs of asymptomatic subjects (Teo et al., 2021).

The lower detection of viral RNA in the saliva of convalescent subjects compared with NP swabs could reflect the lower level of active viral replication in the late phase of the infection. It has been observed that the majority of NP swabs derived from clinically resolved infections, though positive, do not harbour virions that can be isolated in culture. Detection of residual non-replicative RNA in the nasopharyngeal epithelium rather than the saliva, could therefore make the latter a more selective indicator of the presence of infectious virus.

Interestingly, all discordant subjects with negative saliva had low viral loads in the paired nasopharyngeal swabs (Ct ranging between 30 to 40), a result that could be expected for weakly positive samples with viral contents close to the limit of detection of the test. However, some individuals that tested negative in the NP swab method harboured particularly high viral loads in the saliva (Ct ranging between 21 to 26). This discrepancy was not associated with a specific group of samples and likely mirrors the ability of the healthcare operator to collect a successful NP swab, rather than reflecting an inherent property of the biological sample. This is an indication that the saliva test is less prone to variability and therefore more reliable.

In consclusion, given that droplets and aerosol created by saliva are the main transmission route (Zhou et al., 2021), it is reasonable to postulate that individuals that shed high amount of virus in the saliva are also the most contagious and the most crucial to identify. By showing the efficacy and reliability of saliva testing in asymptomatic individuals, our study therefore supports the use of saliva for widespread COVID-19 testing in the community.

## Methods

### Study population

Subjects were recruited at a drive-through testing facility between March 25 and April 23, 2021. The cohort included individuals from the general population undergoing COVID-19 testing for difference reasons. Some individuals were tested because they presented with respiratory symptoms, other had already been diagnosed with COVID-19 and they were tested to confirm their recovery, other were tested following contact tracing and finally a group of health care operators were undergoing regular routine screening. (Table 1). The study received prior approval by the Ethical Committee of the Azienda Provinciale per i Servizi Sanitari of the autonomous province of Trento. An informed consent was obtained from all participants undergoing testing.

### Samples collection

All subjects enrolled underwent testing of NP swabs and saliva. While waiting in line for collection of the NP swab by a healthcare professional, a saliva collection kit (Comedical, Trento, Italy) was distributed to the participants. They collected the saliva autonomously after reading or watching the instructions linked with the kit (https://www.covidsaliva.it/). Inclusion in the study required not having eaten, drunk or smoked for at least 30 minutes prior to saliva collection. The NP swabs were acquired immediately after saliva samples were collected. NP samples were acquired over different days by different healthcare professionals. Both saliva and NP swabs were transported to the laboratory the same day, stored at 4°C overnight and processed for analysis the following day.

### The analytical process

All liquid handling was performed with robotized multichannel pipettes with adjustable tip spacing (Assist Plus, Integra). Saliva samples were first fluidified with the addition of 1 mL DTT 6.5 mM in water directly into the collection tubes followed by pipetting 10 times for resuspending the reducing solution. After this operation the saliva was sufficiently fluidified for further processing. Saliva and NP samples were therefore transferred to 96 deepwell plates (Thermofisher) and extraction performed using the MagMAX™ Viral/Pathogen Nucleic Acid Isolation Kit (Thermofisher) following the manufacturer instruction. After addition of the extraction mix, proteinase K was also added, as indicated by the manufacturer instructions, a step that was found to be crucial for the efficiency of the extraction process. Extraction was operated with a KingFisher™ Flex Purification System (ThermoFisher,) automatic extractor, and RNA was eluted in 50 µl of RNAse-free water. PCR was set-up on 96-well plates using a dedicated robotized multichannel pipette with adjustable tip spacing (Assist Plus, Integra). RT-PCR amplification was performed on CFX96 termocyclers (BioRad) using the Novel Coronavirus Real Time Multiplex RT-PCR Kit (Liferiver) following the manufacturer instructions. Amplification results were analyzed using CFX Maestro software with the single threshold method and Ct calculated by placing the threshold just above the signal given by the molecular grade water negative control sample. Diagnosis was based on the detection of RdRP gene within a threshold value of 40. The kit provides an internal extraction control to be added to each sample, which was used to monitor the efficiency of extraction. A custom-made middleware (Olos, Stardata) was implemented to track the samples through the process, to guide the operators through the analytical phases, to retrieve the amplification results and to communicate the data to the healthcare service repository.

### Statistical analysis

Quantification of agreement (concordance) by kappa and the Pearson correlation coefficient were both calculated using the calculator from GraphPad (Prism).

## Data Availability

The data set analyzed in this paper is available from the corresponding author on request.

## Footnotes

We would like to thank Silvia Contini and Stefania Tissot (APSP Margherita Grazioli) and Lucio Piffer (RSA Volano) for their precious help.

This work was supported by funding from PAT (Provincia Autonoma di Trento) to the University of Trento with a contribution from APSS (Azienda Provinciale per i Servizi Sanitari, Trento) and from the University of Trento.

